# Home confinement during COVID-19 pandemic reduced physical activity but not health-related quality of life in previously active older women

**DOI:** 10.1101/2020.12.21.20248662

**Authors:** Vanessa Teixeira do Amaral, Isabela Roque Marçal, Thiago da Cruz Silva, Fernanda Bianchi Souza, Yacco Volpato Munhoz, Pedro Henrique Camprigher Witzler, Matheus Monge Soares Correa, Bianca Fernandes, Emmanuel Gomes Ciolac

## Abstract

**Background:** To investigate the effect of COVID-19 home confinement on levels of physical activity, sedentary behavior and health-related quality of life (HRQL) in older women previously participating in exercise and educational programs.

**Methods:** 64 older women (age = 72±5 ys) who participated in a physical exercise/educational program and adhered to government home confinement recommendations have their levels of physical activity, sedentary behavior and HRQL assessed before and during (11 to 13 weeks after introduction of government recommendations to reduce virus transmission) COVID-19 pandemic.

**Results:** There were significant reductions in total physical activity (−259 METs/week, *P* = 0.02), as a result of a ∼17.0 % reduction in walking (−30.8 min/week, *P* = 0.004) and ∼41.8 % reduction in vigorous-intensity activity (−29.6 min/week, *P* < 0.001). Sedentary behavior also increased (2.24 h/week, *P* < 0.001; 1.07 h/week days, *P* < 0.001; and 1.54 h/weekend days, *P* < 0.001). However, no significant change occurred in moderate-intensity physical activity, and HRQL domains and facets, except for an improvement in environment domain.

**Conclusion:** Home confinement due to COVID-19 pandemic decreased levels of physical activity and increased levels of sedentary behavior in previously active older women who participated in an educational program. However, there were no significant changes in HRQL. These results suggest that educational programs promoting healthy behaviors may attenuate the impact of home confinement in older women.

## 1. Introduction

The outbreak of coronavirus disease 2019 (COVID-19) is a global public health emergency, responsible for 75,479,471 confirmed cases and 1,686,267 deaths worldwide (data as of December 21, 2020)^1^. Older individuals and those with diabetics, hypertension, and cardiovascular and respiratory diseases are at higher risk of COVID-19 complications and mortality ^2^.

Although the measures to contain the spread of the virus (i.e.: social distancing, home confinement, quarantine and/or lockdown) are effective for reducing incidence and mortality of COVID-19^3^, they may result in negative behaviors that have important repercussions for both individual health and global burden of noncommunicable chronic disease (NCDs). For example, the adoption of these measures has been associated with reduced levels of physical activity ^4,5^, increased levels of sedentary behavior ^4,5^, and a more unhealthy food consumption^4^. Few weeks of these negative behaviors may result in several deleterious cardiometabolic and functional consequences that impact management of NCDs ^6^, mainly in older and high-risk populations ^6–8^.

However, little is known about the effect of measures to contain COVID-19 spread on physical activity levels and health-related quality life (HRQL) of older individuals that are physically active, particularly those who had their group exercise routine interrupted, which may also result in social relation impairments. Thus, our aim was to investigate the effect of COVID-19 home confinement on levels of physical activity, sedentary behavior and HRQL in older women that were previously participating in community-based exercise programs. We hypothesized that home confinement would result in impairments on physical activity, sedentary behavior and HRQL.

## 2. Methods

### 2.1 Population and Study Design

The present study is a longitudinal, mixed-methods, observational study, assessing older women (age > 65 years) that were previously participating in the *Ativa Melhor Idade* project. The *Ativa Melhor Idade* is a university extension program (São Paulo State University [UNESP] at Bauru) that promotes twice-weekly community-based exercise programs and a health-related educational program (i.e.: importance and how to meet the recommended levels of physical activity, to avoid sedentary behavior, and to have good nutritional habits). Participants have their levels of physical activity, sedentary behavior and HRQL assessed before (between January and February 2020) and 11 to 13 weeks after introduction of societal shutdown, self-isolation and social-distancing measures to reduce virus transmission (June 2020) by State of Sao Paulo (Brazil) government. Inclusion criteria included (1) age > 65 ys, (2) regular participation (at least 60% of compliance) in the *Ativa Melhor Idade* project for a minimum of six months prior inclusion in the study, (3) to have responded the physical activity/sedentary behavior and HRQL questionnaires during the project annual assessment before beginning of COVID-19 pandemic, and (4) to adhere the government home confinement recommendations to reduce virus transmission. Participants with cognitive impairment who were unable to answer the questionnaires accurately were not included. Participants who did not answer any questionnaire in the follow-up assessment were excluded. A total of 130 older women volunteered to participate in the study, ninety women met the inclusion criteria and were included in the study. However, only 64 women (age = 72±5 ys; body mass index = 29.0±5.2 kg/m^2^) responded the questionnaires during follow-up assessment and were included in final analysis (Figure 1).

**Figure 1.**
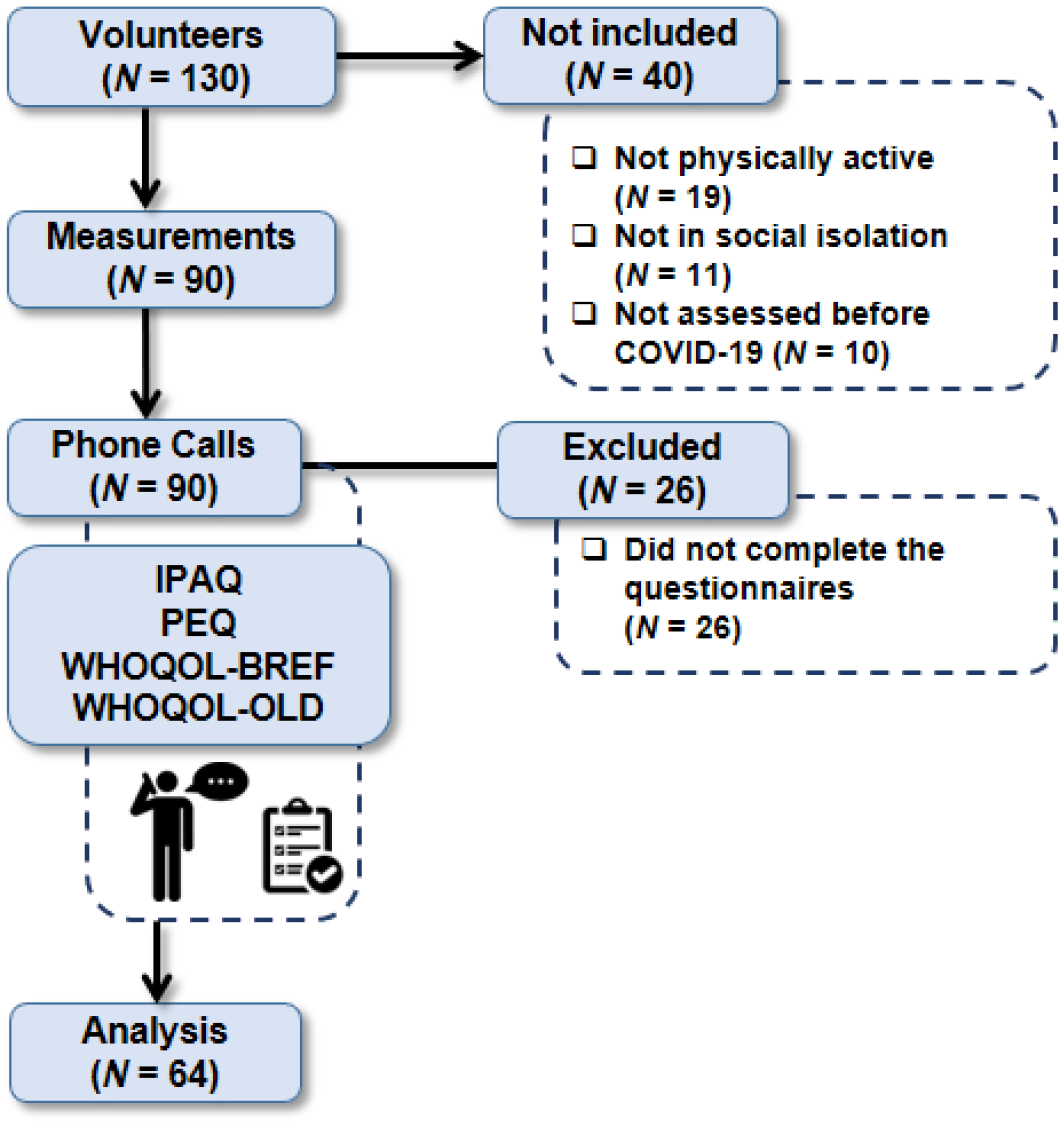
Study design.

Baseline assessment were performed in the Exercise and Chronic Disease Research Laboratory at UNESP (Bauru–SP, Brazil), during the annual assessment of *Ativa Melhor Idade* project (between January and February 2020), and included anamnesis (health status and medications used), and measures of physical activity/sedentary behavior (questionnaire), HRQL (questionnaires), anthropometrics (weight, height and body mass index [BMI]), hemodynamics (blood pressure, heart rate and arterial stiffness), functional levels (flexibility, balance, muscle strength and endurance performance). Follow-up assessment was performed by a telephone call interview (average duration of 50 min), and included anamnesis (health and behavioral status, medications used, compliance to social isolation/home confinement measures, and participants’ perception about the effects of pandemic) and measures of physical activity/sedentary behavior and HRQL. All procedures were approved by the Research Ethics Committee of School of Sciences of UNESP (CAEE 21220919.0.0000.5398) and all participants provided written informed consent.

### 2.2 Measurements

Baseline and follow-up assessments were performed by trained evaluators, during the afternoon. Anamnesis and questionnaires were applied in the following sequence: (1) anamnesis and physical evaluations (only at baseline) to characterize the sample profile; (2) levels of physical activity and sedentary behavior (International Physical Activity Questionnaire [IPAQ] short version) ^9^; (3) a questionnaire assessing compliance to home confinement and to other preventive measures of virus spread (only during follow-up); (4) a questionnaire assessing participants’ perception about the effects of pandemic on daily living activities, health and anxiety, sleep quality, pain, medication use, eating habits, weight, and alcohol and cigarettes consumption (only during follow-up); and (5) HRQL questionnaires (WHOQOL-BREF and WHOQOL-OLD)^10,11^. Weekly time and metabolic equivalents (METs) ^12^ levels of physical activity, as well as the total, domains and facets scores of HRQL were compared between baseline and follow-up.

### 2.3. Statistical Analysis

The Shapiro-Wilk and Levene’s tests were applied to test normality and homoscedasticity of the data assessed before and during COVID-19 pandemic (IPAQ and WHOQOL). Paired samples t-test and Wilcoxon signed-rank test were used to analyze the differences between pre and during COVID-19 variables in parametric and non-parametric data, respectively. Data are presented in mean ± standard deviation (SD), median (interquartile range) or median (minimum–maximum). Participants’ perception about the effects of COVID-19 pandemic are presented descriptively. Statistical software SPSS 17.0™ (SPSS Inc., Chicago, IL, USA) was used to perform the statistical analyses. The statistically significant level was considered when *P* < 0.05.

## 3. Results

All participants had at least one comorbidity (hypertension: *N* = 44; musculoskeletal disorders: *N* = 28; type 2 diabetes Mellitus: *N* = 23; dyslipidemia: *N* = 13; thyroid diseases: *N* = 6; and heart diseases: *N* = 6) and were ∼81 days in social isolation/home confinement. During the pandemic, all participants reported to leave their homes once a week or once every two weeks to buy food and/or medications. Participants reported several behavioral alterations as a consequence of the social isolation/home confinement, including sleeping quality (24 participants reported that their sleep worsened, and 28 participants reported to wake up later than normal), and eating habits (22 participants reported worsening of eating habits, 31 participants reported weight gain and 11 participants reported weight loss). Among the participants who consumed alcoholic beverages (15 participants), three participants reported decreasing weekly alcohol consumption, and 12 participants reported no change. Among smokers (10 participants), two participants reported an increase in cigarette consumption, and eight participants reported no change. Sixty participants reported to have some kind of pain in their body, where 15 participants reported an increase and 11 participants reported a decrease during COVID-19 pandemic. Fifty-eight participants reported to maintain routine medication consumption, and six participants reported an increase (*N* = 3) or decrease (*N* = 3) in routine medication consumption.

Participants also reported that COVID-19 pandemic affected very much or completely their physical activity routine (*N* = 44). Fifty-one participants reported that were exercising regularly during this period, where 44 participants were exercising at home and seven participants were exercising outdoor (public spaces). Among those who were exercising, 29 participants reported to receive professional guidance remotely. The pre *vs*. during COVID-19 pandemic assessment showed significant reductions in total physical activity (−259 METs/week, Z = −2.32, *P* = 0.02), as a result of a ∼17.0 % reduction in walking (−30.8 min/week; Z = −2.90, *P* = 0.004) and ∼41.8 % reduction in vigorous-intensity activity (−29.6 min/week, Z = −3.93, *P* < 0.001); however, no significant change occurred in moderate-intensity activity (Figure 2). Sedentary behavior (total sitting time) also increased (2.24 h/week, Z = −3.70, *P* < 0.001) during COVID-19 pandemic in both week days (1.07 h/week, Z = −3.64, *P* < 0.001) and weekend days (1.54 h/week, Z = −3.48, *P* < 0.001) (Figure 2).

**Figure 2.**
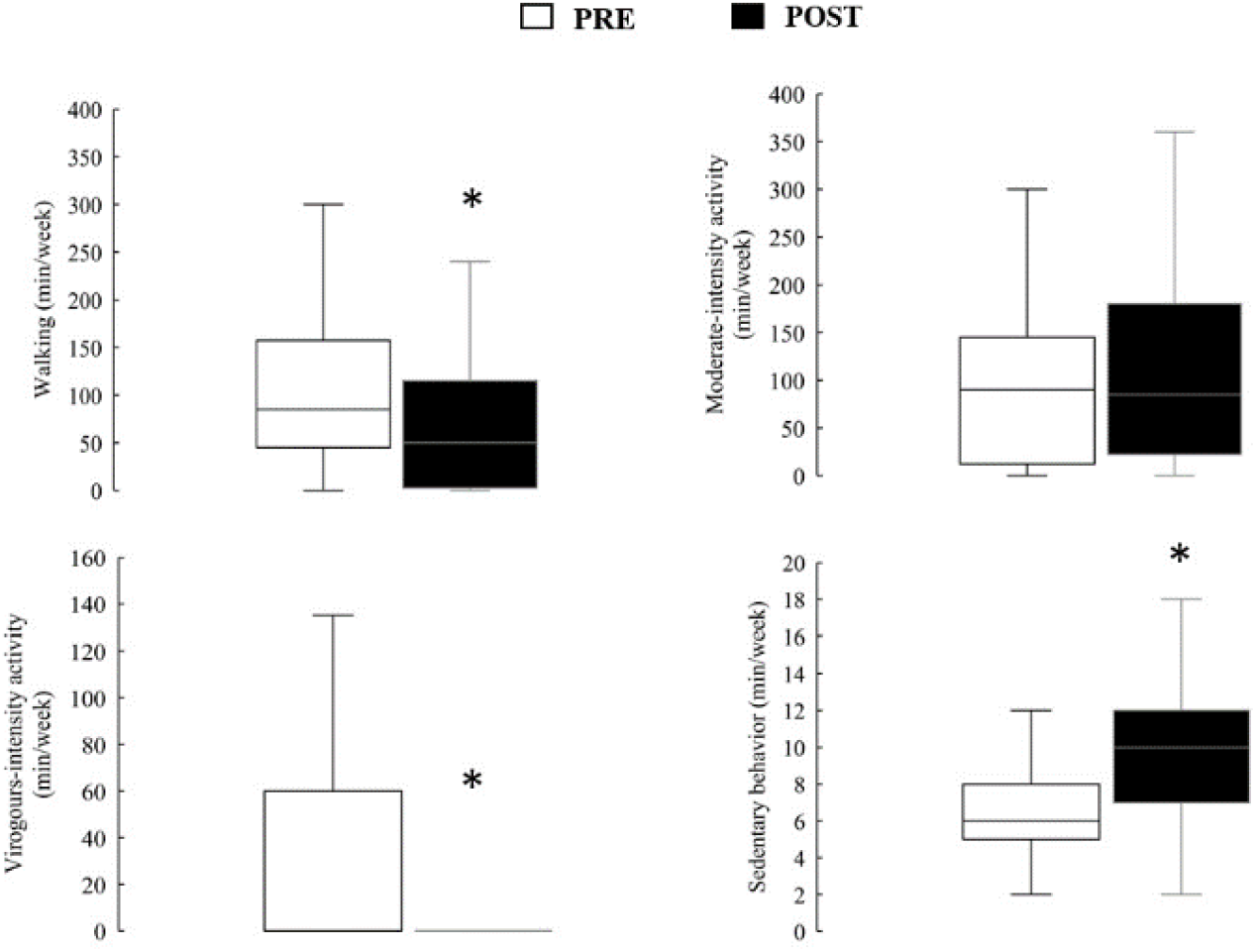
Levels of physical activity and sedentary behavior before and during home confinement. *: significant difference between pre and post (*P* < 0.05).

However, HRQL domains and facets did not change during COVID-19 pandemic, except for an improvement in environment domain (Table 1).

**Table 1.** Quality of life before and during home confinement/quarantine.

| WHOQOL-BREF |  |  |  |
| --- | --- | --- | --- |
| Domains | PRE | POST | *P* |
| Physical Health | 14.3 ± 2.5 | 14.4 ± 2.8 | 0.61 |
| Psychological | 15.4 ± 2.2 | 15.6 ± 2.5 | 0.39 |
| Social Relationship | 15.7 ± 2.7 | 15.3 ± 2.7 | 0.25 |
| Environment | 14.0 ± 2.2 | 14.6 ± 2.2 | 0.01 * |
| Global QOL | 15.2 ± 2.9 | 15.5 ± 2.9 | 0.38 |
| WHOQOL-OLD |  |  |  |
| Facets | PRE | POST | *P* |
| Sensory Abilities | 14.8 ± 3.2 | 15.5 ± 3.2 | 0.09 |
| Autonomy | 14.1 ± 2.9 | 14.6 ± 3.1 | 0.13 |
| Past, Present and Future Activities | 14.5 ± 2.7 | 14.9 ± 2.1 | 0.34 |
| Social Participation | 15.4 ± 2.8 | 14.7 ± 2.7 | 0.09 |
| Death and Dying | 14.3 ± 4.1 | 14.1 ± 4.4 | 0.67 |
| Intimacy | 14.7 ± 3.5 | 15.6 ± 3.6 | 0.08 |

\*: Significant difference between pre and post.

## 4. Discussion

The main findings of present study were that older women who were physically active prior to COVID-19 pandemic showed substantial reduction in physical activity and increase in sedentary behavior during the first months of home confinement. However, there were no significant changes in HRQL, except for a small improvement in environment domain. According to our knowledge, this is the first longitudinal study that investigated the effect of COVID-19 home confinement on physical activity, sedentary behavior and HRQL in older women that were physically active prior to pandemic.

Cross-sectional studies has shown that social isolation and home confinement imposed during COVID-19 pandemic reduced the vigorous activities and walking in reduced levels of physical activity and increased levels of sedentary behavior in individuals aged < 65 ys ^4,13^ In addition, a small qualitative study also suggested that home confinement affected the number of older people who participated in group physical activity programs^14^. The reduced levels of physical activity and increased levels of sedentary behavior in the present study, as well as the high prevalence (69 %) of participants reporting that COVID-19 pandemic affected very much or completely their physical activity routine study, is in accordance with previous cross-sectional ^4,13^ and qualitative ^14^ studies. However, it is important to note that the reduction in walking found in the present study (17.1 %) was lower than the reductions in the previous studies with participants aged < 65 ys (34 % to 58.2 %) ^4,13^. In addition, the previous cross-sectional studies showed reductions in substantial reductions in moderate-intensity physical activity ^4,13^, which did not change in the present study. Although differences in age of the population and in study design may not be discharged as reasons for the lower physical activity reduction in the present than previous studies ^4,13^, the fact that participants of present study were physically active and participated in an educational program to promote healthy behaviors prior to pandemic should be considered. Educational programs promoting healthy behaviors has shown to be effective in improving physical activity ^15,16^. Thus, it is possible that the participation in the educational program prior to COVID-19 pandemic may be attenuated the walking and moderate-intensity activity reduction in the present study. Future studies are welcome to confirm this hypothesis.

The positive effects of physical activity on HRQL in older individuals are well known ^17,18^. In this context, we hypothesized that home confinement due to COVID-19 pandemic would result in reduction of physical activity and, consequently, HRQL in participants of present. However, despite reduction in physical activity, there were no significant changes in 10 out of 11 domains/facets of HRQL. The small reduction in physical activity and the prior participation in the educational program may be associated with the no change in HRQL in the present study. It is important to note that, in contrast to our hypothesis, there was a significant increase in environment domain of HRQL during follow-up. This improvement may have occurred because participants felt more secure and protected at home during a pandemic. In addition, as participants of present study were retired women who have no change in retirement salary, it is possible that home confinement resulted in greater savings in financial resources, affecting positively HRQL environment domain during follow-up.

The lack of a comparison group with older individuals who were not participating in physical activity programs and/or an educational program prior to COVID-19 pandemic is a limitation of present study, because it does not allow us to affirm that the present small reduction in physical activity and the no change in HRQL was due to prior physical exercise and/or educational program participation. The measure of physical activity levels by questionnaire, instead of more accurate tools such as accelerometers is also a limitation that should be acknowledged.

The present study has also important implications. Low levels of physical activity is associated with innumerous age-related cardiometabolic and functional impairments ^6,7,19^. For example, few weeks of reduction in levels of physical activity may result in deleterious effects on glycemic control, body composition, inflammatory cytokines, blood pressure, vascular function, cardiorespiratory/muscle fitness, balance and agility ^6,7,19^. In addition, reductions in levels of physical activity, as the observed in the present study, may result in increased incidence of NCDs and mortality if persisting for long periods ^6^. Finally, it is also important to note that we found substantial increases in the levels of sedentary behavior, which may potentiate the above-mentioned deleterious effects of reduced levels of physical activity ^6^. In this context, strategies for promoting adequate levels of physical activity and reducing sedentary behavior during COVID-19 home confinement should be urgently implemented.

## 5. Conclusion

In summary, home confinement due to COVID-19 pandemic resulted in decreased levels of physical activity and increased levels of sedentary behavior in previously active older women who participate in an educational program promoting healthy behaviors. However, there were no significant changes in HRQL. As the reduction in physical activity was lower in the present study than in previous cross-sectional studies ^4,13^, it can be suggested that educational programs promoting healthy behaviors may attenuate the impact of home confinement in physical activity. Future studies addressing this hypothesis are welcome.

## Data Availability

The data used to support the findings of this study are available from the corresponding author upon request.

## Acknowledgements

VTA and BF were supported by *Coordenação de Aperfeiçoamento de Pessoal de Nível Superior (CAPES)*. IRM and EGC were supported by *Fundação de Amparo à Pesquisa do Estado de São Paulo* (FAPESP # 2018/09695-5) and *Conselho Nacional de Desenvolvimento Científico e Tecnológico* (CNPq #303399/2018-0), respectively. The results of the present study are presented clearly, honestly, and without fabrication, falsification, or inappropriate data manipulation.

## Author’s Contributions

VTA participated in study design, literature review, data collection and analysis, and manuscript writing. TCS, FBS, YVM, PHCW and MMSC participated in data collection and manuscript writing. IRM and BF participated in data analysis and manuscript writing. EGC prepared the study, supervised the division of tasks, and participate in data analysis and manuscript writing. All authors approved the final version of the manuscript.

## References

1. World Health Organization. Coronavirus disease (COVID-19). Accessed November 16, 2020. https://www.who.int/emergencies/diseases/novel-coronavirus-2019

2. Jiménez-Pavón, D, Carbonell-Baeza, A, Lavie, C J. Physical exercise as therapy to fight against the mental and physical consequences of COVID-19 quarantine: Special focus in older people. Prog Cardiovasc Dis. Published online 2020:386-388. doi:https://doi.org/10.1016/j.pcad.2020.03.009

3. Nussbaumer-Streit B, Mayr V, Dobrescu AI, et al. Quarantine alone or in combination with other public health measures to control COVID-19: a rapid review. Cochrane Database Syst Rev. 2020;2020(9). doi:10.1002/14651858.CD013574.pub2

4. Ammar A, Brach M, Trabelsi K, et al. Effects of COVID-19 Home Confinement on Eating Behaviour and Physical Activity: Results of the ECLB-COVID19 International Online Survey. Nutrients. 2020;12(6):1583. doi:10.3390/nu12061583

5. Visser M, Schaap LA, Wijnhoven HAH. Self-Reported Impact of the COVID-19 Pandemic on Nutrition and Physical Activity Behaviour in Dutch Older Adults Living Independently. Nutrients. 2020;12. doi:10.3390/nu12123708

6. Marçal IR, Fernandes B, Viana AA, Ciolac EG. The Urgent Need for Recommending Physical Activity for the Management of Diabetes During and Beyond COVID-19 Outbreak. Front Endocrinol (Lausanne). 2020;11. doi:10.3389/fendo.2020.584642

7. Ciolac EG, Rodrigues da Silva JM, Vieira RP. Physical Exercise as an Immunomodulator of Chronic Diseases in Aging. J Phys Act Heal. 2020;17(6):662–672. doi:10.1123/jpah.2019-0237

8. McGlory C, von Allmen MT, Stokes T, et al. Failed Recovery of Glycemic Control and Myofibrillar Protein Synthesis With 2 wk of Physical Inactivity in Overweight, Prediabetic Older Adults. Journals Gerontol Ser A. 2018;73(8):1070–1077. doi:10.1093/gerona/glx203

9. Questionnaire G for data processing and analysis of the IPA, Forms (IPAQ)—Short and Long. Guidelines for data processing and analysis of the International Physical Activity Questionnaire (IPAQ)-short and long forms. IPAQ Web site. Published 2005. Accessed December, 21, 2020. https://sites.google.com/site/theipaq/scoring-protocol

10. The Whoqol Group. Development of the World Health Organization WHOQOL-BREF Quality of Life Assessment. Psychol Med. 1998;28(3):551–558. doi:10.1017/S0033291798006667

11. Power M, Quinn K, Schmidt S. Development of the WHOQOL-Old module. Springer. 2005;14(10):2197–2214. doi:10.1007/s11136-005-7380-9

12. Pardini R, Matsudo S, Araújo T, et al. Validation of the international questionnaire of physical activity level (IPAQ—version 6): pilot study in young Brazilian adults. Rev Bras Ciên e Mov Brasília. 2001;9:45–51.

13. Castañeda-Babarro A, Arbillaga-Etxarri A, Gutiérrez-Santamaria B, Coca A. Impact of COVID-19 confinement on the time and intensity of physical activity in the Spanish population. Research Square. Published online 2020. doi:10.21203/rs.3.rs-26074/v1

14. Goethals L, Barth N, Guyot J, Hupin D, Celarier T, Bongue B. Impact of Home Quarantine on Physical Activity Among Older Adults Living at Home During the COVID-19 Pandemic: Qualitative Interview Study. JMIR Aging. 2020;3(1):e19007. doi:10.2196/19007

15. Da Silva JMR, De Rezende MU, Spada TC, et al. Educational program promoting regular physical exercise improves functional capacity and daily living physical activity in subjects with knee osteoarthritis. BMC Musculoskelet Disord. 2017;18(1). doi:10.1186/s12891-017-1912-7

16. Da Silva JMR, De Rezende MU, Spada TC, et al. Muscular and functional capacity in subjects under treatment for knee osteoarthritis: Role of physical activity status. J Phys Act Heal. 2019;16(5):362–367. doi:10.1123/jpah.2018-0318

17. Marquez DX, Aguiñaga S, Vásquez PM, et al. A systematic review of physical activity and quality of life and well-being. Transl Behav Med. 2020;10(5):1098–1109. doi:10.1093/tbm/ibz198

18. Scarabottolo CC, Cyrino ES, Nakamura PM, et al. Relationship of different domains of physical activity practice with health-related quality of life among community-dwelling older people: A cross-sectional study. BMJ Open. 2019;9(6):1–10. doi:10.1136/bmjopen-2018-027751

19. Ciolac EG. Exercise training as a preventive tool for age-related disorders: A brief review. Clinics. 2013;68(5):710–717. doi:10.6061/clinics/2013(05)20

